# Exploring quantitative MRI biomarkers of head and neck post-radiation lymphedema and fibrosis: Post hoc analysis of a prospective trial

**DOI:** 10.1101/2024.06.30.24309685

**Authors:** MD Anderson Head and Neck Cancer Symptom Working Group, Shitong Mao, Jihong Wang, Holly McMillan, Abdallah Sherif Radwan Mohamed, Sheila Buoy, Sara Ahmed, Samuel L Mulder, Mohamed A. Naser, Renjie He, Kareem A. Wahid, Melissa Chen, Yao Ding, Amy C. Moreno, Stephen Y. Lai, Clifton D. Fuller, Katherine A. Hutcheson

## Abstract

**Importance:** Quantifying Head and Neck Lymphedema and Fibrosis (HN-LEF) is crucial in the investigation and management of this highly prevalent treatment sequelae in head and neck cancer (HNC). The HN-LEF grading system classifies physically palpable soft-tissue injury categorically. Imaging biomarkers from MRI may serve to complement or validate physical HN-LEF grading when assessing the effectiveness of therapeutic interventions or toxicity profiles of patients.

**Objective:** To explore the relationship between 1) physical HN-LEF classification in submental and oral regions of interest (ROI) and the MRI T1- and T2-weighted signal intensity (SI) in close proximity regions, and 2) a novel HN-LEF score and MRI T1 and T2 structural volumes.

**Design:** Post hoc analysis of pilot single arm MANTLE trial (NCT03612531).

**Setting:** Single institution, NCI-designated comprehensive cancer center.

**Participants:** A total of 16 individuals (mean [SD] age, 68.28 [7.0] years; 3[19%] female) enrolled in the MANTLE trial underwent MRI. All participants were disease-free at least two years post radiotherapy with grade ≥2 fibrosis (in any cervical ROI) and grade ≥2 dysphagia (per DIGEST). Over a 12-week period, participants engaged in manual therapy sessions accompanied by concurrent standardized multiparametric, serial MRI examinations and palpation-based HN-LEF evaluations at 3 time points: baseline, post-manual therapy, and post-washout.

**Exposures:** The independent variable HN-LEF included its categorical classification (No-LEF, A-B = edema, C= edema + fibrosis, D=fibrosis) and a novel metric (10-point scale) derived from the HN-LEF categories (considering both type and severity classification).

**Main Outcomes and Measures:** The T1- and T2-weighted MRI SI was examined by Kruskal-Wallis tests in relation to HN-LEF categories and the novel HN-LEF score. We hypothesized higher T2 SI in edema states, higher T1 SI in fibrotic states, and decreasing structural volume as HN-LEF score increased.

**Results:** We identified differences in mean ranks among HN-LEF categories in relation to the MRI SI (A-B and C are higher than D and No-LEF for T2 SI, and A-B is the highest for T1). Furthermore, six pairs of FOM volumes on MRI demonstrated a strong negative correlation (p<0.05) with the HN-LEF score at adjacent palpable sites: digastric vs. submental left (*ρ* = -0.421; 95% CI, -0.65∼ -0.10, T1), mylohyoid vs. submental left (*ρ* = -0.36; 95% CI, -0. 62∼ 0.03, T1), digastric vs. submental left (ρ = --0.45; 95% CI, -0. 72∼ -0.06, T2), genioglossus vs. Intraoral left (ρ = -0.47; 95% CI, -0. 74∼ -0.07, T2), mylohyoid vs. Intraoral left (ρ = -0.48; 95% CI, -0. 75∼ -0.09, T2), tongue base vs. Intraoral left (r = -0.42; 95% CI, -0. 71∼ -0.01, T2).

**Conclusions and Relevance:** This exploratory analysis provides hypothesis generating data supporting further study of MRI SI as an imaging biomarker of edematous soft tissue states after RT in HNC, but does not support the hypothesized T2 SI relationship with fibrotic tissue states. The inverse correlation between the novel HN-LEF scores and structural volumes points to the potential validity of this novel metric assuming structural volume diminishes as patients move from edema to fibrotic states. This study highlights the potential for enhancing the LEF quantification using imaging metrics, which might further aid in the early detection and precise measurement of lymphedema and fibrosis severity in post-radiation HNC patients.

## 1. Introduction

Advancements in multimodal treatments for head and neck cancer (HNC), including surgery, radiotherapy (RT), and chemotherapy, have significantly improved patient survival rates(1). However, these comprehensive treatments have also increased the incidence of head and neck Lymphedema and Fibrosis (HN-LEF), which are major contributors to post-treatment functional impairments in HNC survivors, affecting up to 90% of patients(2). RT fibrosis is particularly challenging, leading to tissue stiffening that impacts aesthetics and functions such as swallowing, speaking, and cervical range of motion (3-5). Intraoral lymphedema can cause swelling of oral tissues, impairing speech clarity, chewing efficiency, and swallowing safety, thus increasing the risk of aspiration. Fibrosis can also lead to trismus, restricting mouth opening and complicating oral hygiene and nutrition, thereby significantly affecting quality of life(6). Early identification and timely referral to certified therapists are crucial for effective management of LEF. Furthermore, the variability in RT techniques results in diverse LEF manifestations, underscoring the importance of advancing LEF assessments to personalize interventions for each patient.

Quantitative assessment of LEF in the oral and submental regions among HNC patients following RT presents considerable challenges(8, 9). Currently, two main methodologies exist: clinician-graded evaluations and technical/imaging-based evaluations(10). Clinician-reported assessments, based on visual inspection and palpation, are commonly utilized due to their inexpensive, practical, and accessible features (11). However, clinically validated tools for palpation-based grading remain scarce. Deng et al. developed an innovative LEF grading framework combining nominal (type classification) and ordinal (severity grading) variables(6, 8, 12), which demonstrated strong inter-rater and intra-rater reliability. While clinical assessments are valuable, they are subjective, lack sensitivity to smaller degrees of change that may be clinically meaningful, and have not been benchmarked to objective, quantifiable data from technical/imaging-based evaluations.

Magnetic Resonance Imaging (MRI) is a promising tool to evaluate the impact of LEF on HN muscles, particularly the floor of mouth (FOM) muscles. MRI imaging allows visualization of tissue composition and structure, distinguishing between normal tissue, edematous changes, and the dense, scarred tissue. T1-weighted MRI sequences are particularly effective for identifying fibrotic tissue as fibrosis tends to appear with higher signal intensity (SI) compared to normal muscle tissue (13). T2-weighted images are invaluable for highlighting areas with increased fluid content, as seen in lymphedema, by showing higher SI(14-16). This capability is essential for assessing the severity and extent of LEF, with potentially vital clinical insights into the impact on muscle function. MRI biomarkers have potential to become an essential tool for monitoring the progression of LEF and evaluating therapeutic outcomes, helping to tailor treatment plans to improve patients’ quality of life and functional outcomes.

This study explores the relationship between palpation based LEF measurement and MRI as a potential quantitative imaging biomarker for assessing HN-LEF. We further explore a novel HN-LEF metric from physical measurement. Our previous work introduced a pilot trial of manual therapy (NCT03612531), where both MRI and LEF measures were conducted concurrently to evaluate therapeutic progress in fibrosis-related dysphagia in HNC survivors(17, 18). Building on that foundation, this research targets two critical connections: A) the alignment between HN-LEF classification of physically palpable soft-tissue variation and quantitative MRI information, alongside B) the link between our novel HN-LEF metric and MRI-delineated structural volumes. These explorations aim to advance therapeutic assessments, optimize patient care protocols based on toxicity profiles, and improve the quality of life for HNC survivors. This endeavor exemplifies the synergy between palpation-based evaluations and advanced imaging techniques.

## 2. Methods

### 2.1 Participants and data collection

This research was conducted as a post hoc analysis of the MANTLE trial (Manual Therapy in Treating Fibrosis-Related Late Effect Dysphagia in Head and Neck Cancer Survivors, NCT03612531), a prospective single-arm unblinded pilot trial of MT in post-RT patients with late dysphagia(17, 18). The trial was undertaken at the University of Texas, MD Anderson Cancer Center (Houston, Texas, USA), an NCI-recognized comprehensive cancer center. The research protocol was approved by MD Anderson Institutional Review Board (IRB: 2018-0052).

The study’s cohort was derived from individuals previously enrolled in the MANTLE trial. All patients signed informed consent, including permission to review medical records, before initiating any study activities. Eligibility was determined based on the following criteria: 1). Disease-free adult participants (age 18 or older); 2). Underwent curative-intent RT for HNC at least two years before the study; 3). Late Dynamic Imaging Grade of Swallowing Toxicity (DIGEST, version 2) grade of 2 or higher, indicating at least moderate grade dysphagia(19, 20); 4). Grade 2 or higher fibrosis, according to CTCAE v4.0 standards (21); 5). Ability and willingness to attend all therapy sessions. Exclusion criteria: 1) Individuals with active recurrent or second primary head and neck, central nervous system, or thoracic cancer at time of enrollment; 2) Presence of active osteoradionecrosis or other non-healing wounds in the manual therapy region of interest, including fistulas, ulcers, or soft tissue necrosis; 3) A history of substantial surgical interventions such as subtotal or total glossectomy or total laryngectomy; 4) Significant chronic or acute cardiac, pulmonary, or neuromuscular diseases that could limit participation; 5) Having a tracheostomy at the time of enrollment.

Among 26 patients consented to the trial, the study cohort consisted of 16 individuals (mean age 68.28 ± 7.0 years; 19% female) who completed MRI (61.5% completion rate). Over a span of 12 weeks, participants were involved in manual therapy sessions aimed at mitigating lymphedema and fibrosis. Assessments were scheduled at three key stages: before the manual therapy sessions began (pre-MT), immediately following the conclusion of the six weeks of manual therapy (post-MT), and after another subsequent six-week period of home-based exercises (post-washout). At each of these stages, a dual-evaluation methodology was employed, which included both standardized multiparametric, serial MRI examinations (including T1- and T2-weighted imaging) and palpation-based HN-LEF evaluations for each participant. The number of T1-weighted imaging was 25(40.3%), and the number of T2-weighted imaging was 37(59.7%).

### 2.2. LEF assessment criteria

To assess LEF, this study utilized the HN-LEF scoring system. This scoring system incorporates both a nominal and an ordinal component to provide a comprehensive evaluation of LEF. The nominal component categorizes LEF into distinct categories (No LEF, A-B, C, D), each representing different stages of LEF. In the first task, where we are assessing whether the SI of specific structures differs across various LEF categories, the independent variable is the LEF categories:

#### Type No LEF

No LEF present. Characterized by the absence of visible tissue swelling and the lack of palpable thickening or tightness in the dermis, suggesting normal tissue integrity without any signs of LEF.

#### Type A-B

Lymphedema alone or dermal thickening. Characterized by visible and palpable soft tissue swelling. The tissues involved are soft when touched, indicating fluid accumulation and the swelling.

#### Type C

Lymphedema and fibrosis. Characterized visible swelling in the soft tissues, but the tissues feel firm to the touch. This indicates a more complex condition where fluid accumulation is accompanied by fibrotic changes, making the swelling non-reducible and persistent.

#### Type D

Fibrosis alone. Characterized by skin and firm tissues, showing increased density and decreased compliance, but without visible swelling.

The ordinal component grades the severity of LEF on a scale from 0 to 5, allowing for a detailed assessment of the progression and intensity of the condition. For the clinical implementation of the LEF assessment, palpation techniques are used by trained clinicians to determine the LEF type and severity, focusing particularly on the submental and intraoral regions:

#### Submental Region (Left and Right)

The submental area, located directly under the chin (mentum), is assessed. This region is between the lower border of the mandible and superior to the hyoid bone, forming the floor of the mouth, externally.

#### Intraoral Region (Left and Right)

Which includes the entire mouth cavity. This area covers the mucosal lining of the cheeks, the FOM (space under the tongue), and the entire palate (maxilla and velum).

### 2.3 Novel HN-LEF scores

In this study, we introduce a novel HN-LEF scoring system that creates a single-value 10-point score (0 to 9) from the palpation-based LEF measurements, and this scoring system is the independent variable for investigating the connection with the structural volumes. This score is derived by cross-referencing the LEF type with its severity on a tabulated matrix, as shown in Table I.

**Table I.** The Conversion Table to derive the Novel HN-LEF Score.

| Novel HN-LEF Score |  | LEF severity |  |  |  |  |  |
| --- | --- | --- | --- | --- | --- | --- | --- |
|  |  | 0 | 1 | 2 | 3 | 4 | 5 |
| LEF types | No-LEF | 0 | 1 | 2 | 3 | 4 | 5 |
|  | A | 1 | 2 | 3 | 4 | 5 | 6 |
|  | B | 2 | 3 | 4 | 5 | 6 | 7 |
|  | C | 3 | 4 | 5 | 6 | 7 | 8 |
|  | D | 4 | 5 | 6 | 7 | 8 | 9 |

### 2.3 MRI imaging and segmentation

In this study, MRI was employed focusing on the precision and detail offered by T1-weighted and T2-weighted images in the head and neck regions. MRI scans were performed using detailed specifications to ensure high-resolution imaging: a slice thickness of 4mm and pixel spacing varying between 0.25mm, 0.28125mm, and 0.3125mm depending on the specific scan sequence used. The MRI data were stored and analyzed using DICOM (Digital Imaging and Communications in Medicine) standards, with specific parameters set for imaging quality and detail resolution: Bits Allocated at 16, Bits Stored at 12, and High Bit at 11. This configuration allowed for a wide dynamic range in signal (voxel) intensity, spanning from 0 to 4096.

The MRI imaging was manually segmented to specifically delineate six anatomical structures relevant to HN-LEF in the submental region: the Digastric anterior (left and right), Genioglossus (oral tongue), Geniohyoid, Mylohyoid, and tongue base. The tongue base refers to the posterior third of the tongue, which is posterior to the circumvallate papillae and proximal to the throat.

The segmentation process was performed by RayStation software (Version 11B DTK, RaySearch Laboratories, Stockholm, Sweden) and resulted in 3D masks corresponding to the targeted tissue areas, which facilitated subsequent quantitative analyses. SI for each area was determined by averaging the voxel intensities within the respective mask. Volume measurements were calculated by summing the voxel counts within each mask and multiplying by the voxel volume (the product of slice thickness and the square of pixel spacing) in cubic centimeters (cm3). This systematic approach allowed for precise quantification of anatomical features and their alterations within the specified regions.

### 2.4 Statistical analysis

Descriptive statistics, including mean, median, standard deviation (STD), and mean of rank of the SI, were summarized for all the structures within each LEF category. The Kruskal-Wallis test was used for overall SI differences among all the LEF categories. Post-hoc analysis using Dunn’s test was conducted to identify specific group differences for different categorical pairs, if the p-value of the Kruskal-Wallis test was lower than 0.05. The Spearman Rank Correlation Coefficient (*ρ*) was used to assess the linear relationship between MRI structure volumes and the novel HN-LEF score, and 95% confidence intervals (CI) were calculated for the correlation coefficients. A *t*-test was used to determine the statistical significance of the Pearson correlation coefficient. Statistical analysis was performed using Python (3.9.16).

## 3. Results

The SI in both the T1- and T2-weighted MRI are summarized in Table II and their distributions by LEF types (ordinal categories) plotted in

**Table II.** Characteristics of MRI SI and Distribution by LEF Categories and MRI Types.

| MRI Sequences |  | T1-Weighted |  |  |  | T2-Weighted |  |  |  |
| --- | --- | --- | --- | --- | --- | --- | --- | --- | --- |
| LEF Categories |  | No-LEF | A-B | C | D | No-LEF | A-B | C | D |
| MRI SI by LEF Categories | Mean | 353.49 | 393.30 | 364.18 | 348.14 | 207.33 | 239.75 | 313.88 | 170.09 |
|  | Median | 377.32 | 414.28 | 235.76 | 344.04 | 189.14 | 224.29 | 238.64 | 150.36 |
|  | STD | 167.9 | 142.48 | 238.08 | 133.23 | 118.55 | 105.22 | 190.25 | 88.70 |
|  | Mean of Rank | 291.36 | 336.71 | 262.05 | 277.38 | 194.87 | 237.11 | 258.40 | 140.16 |
|  | Kruskal-Wallis test p | <0.01 |  |  |  | <0.01 |  |  |  |
|  | Post-hoc analysis | A-B > C (p=0.02), A-B > D (p=0.02), A-B > No-LEF (p=0.03) |  |  |  | A-B > D (p<0.01), A-B > No-LEF (p<0.01), C > D (p<0.01), C > No-LEF (p=0.02), No-LEF > D (p<0.01) |  |  |  |
| MRI SI in FOM Muscles by LEF Categories | Mean | 350.83 | 394.48 | 367.58 | 325.47 | 182.60 | 242.74 | 426.23 | 150.60 |
|  | Median | 371.26 | 419.47 | 206.78 | 306.77 | 172.09 | 224.05 | 414.06 | 150.12 |
|  | STD | 161.39 | 157.39 | 291.30 | 77.32 | 83.28 | 115.14 | 165.63 | 53.81 |
|  | Mean of Rank | 135.55 | 153.07 | 109.68 | 120.44 | 116.3 | 156.96 | 236.83 | 80.44 |
|  | Kruskal-Wallis test p | 0.01 |  |  |  | <0.01 |  |  |  |
|  | Post-hoc analysis | A-B > C (p=0.03) |  |  |  | A-B > D (p<0.01), A-B > No-LEF (p<0.01), C > A-B (p<0.01), C > D (p<0.01), C > No-LEF (p<0.01), No-LEF > D (p=0.02) |  |  |  |
| MRI SI in Intraoral Muscles by LEF Categories | Mean | 368.93 | 420.48 | 419.00 | 315.30 | 184.42 | 178.76 | 132.01 | 139.47 |
|  | Median | 407.67 | 438.07 | 418.59 | 246.05 | 180.62 | 181.56 | 133.97 | 105.73 |
|  | STD | 143.55 | 90.84 | 11.65 | 147.85 | 67.22 | 55.61 | 21.60 | 60.65 |
|  | Mean of Rank | 32.92 | 40.42 | 37.75 | 25.32 | 27.55 | 26.25 | 16.5 | 17.0 |
|  | Kruskal-Wallis test p | 0.12 |  |  |  | 0.11 |  |  |  |

Figure 1. For T1-weighted MRIs, the mean SI for category A-B was consistently higher than for categories C, D, and No-LEF, suggesting that A-B, typically indicative of edema, shows a greater T1 SI compared to other categories. The Kruskal-Wallis tests confirm these differences are statistically significant, with post-hoc analysis indicating that A-B is higher than C, D, and No LEF states. In T2-weighted MRIs, C categories showed highest mean SI followed by A-B compared to D and No-LEF. Counter to hypothesis particularly noting that category C (lymphedema and fibrosis) also exhibited higher T2 SI than the fibrosis-only category D and the No-LEF category.

**Figure 1.**
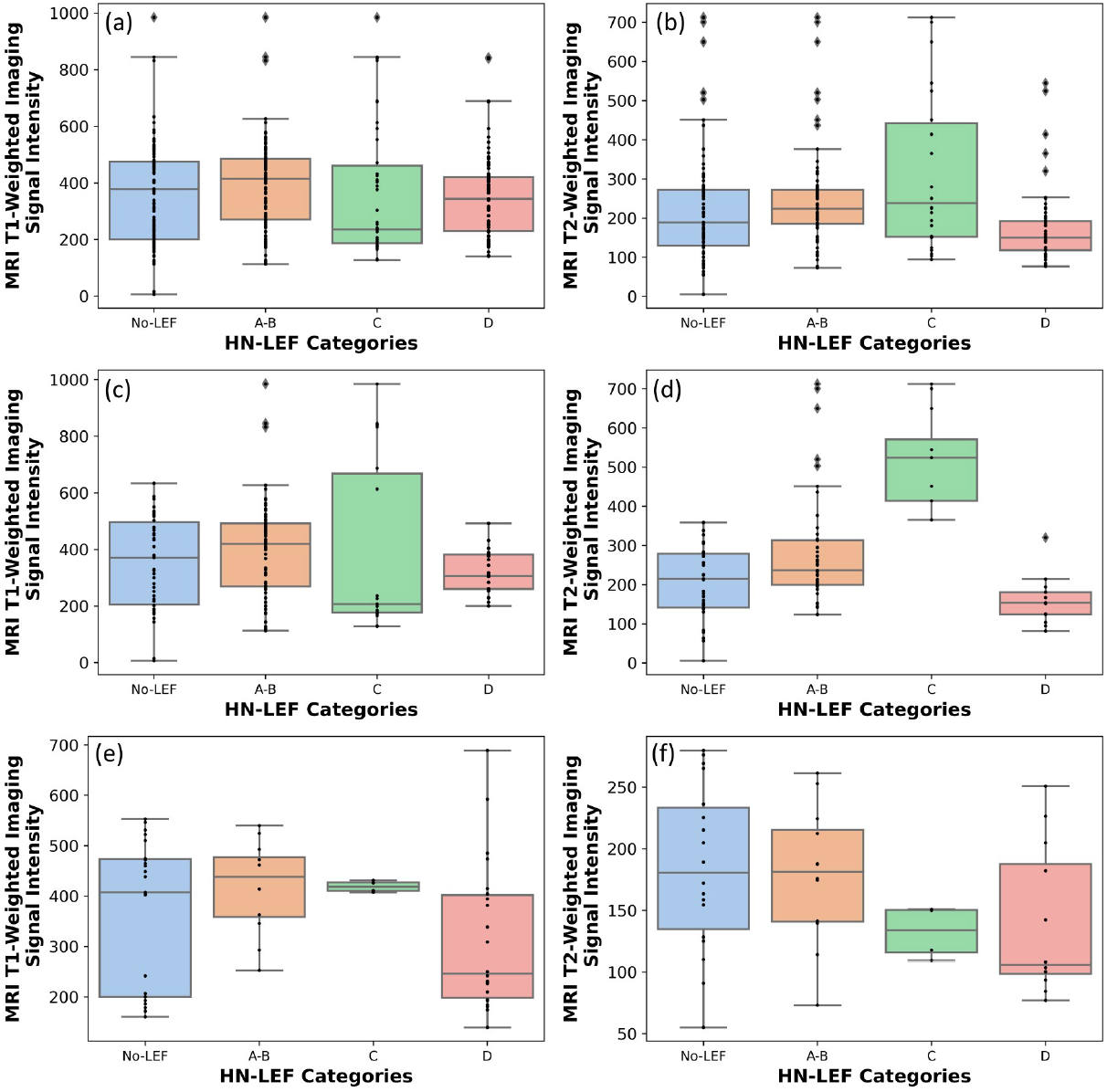
Box plots of the SI distribution. (a), (c), and (e) display T1-weighted MRI data, while panels (b), (d), and (f) show T2-weighted MRI data (a) and (b) present the overall distributions of SI across all palpation sites by LEF categories; (c) and (d) depict the SI distribution of the FOM muscles by LEF categories in the submental area; (e) and (f) illustrate the SI distribution of the tongue oral and tongue base structures by LEF categories in the intraoral area.

Additionally, for specific muscle group, the FOM muscles in submental area, the pattern persisted with A-B and C showing higher SI in T2-weighted images. For the intraoral structures (tongue base and oral tongue), the MRI SI shows varied results across different LEF types.

The correlations between muscle/structure volumes and Novel HN-LEF scores were summarized in Table III. For T1-weighted MRI, the digastric anterior muscle showed a negative correlation in the submental left region (coefficient: -0.41, 95% CI: -0.65 to -0.10). The mylohyoid muscle exhibited a negative correlation in the same palpation site (coefficient: -0.36, 95% CI: -0.62 to 0.03). The Genioglossus, Geniohyoid, Tongue Base, and Oral Tongue muscles did not display any notable correlations with p-values below 0.05. For T2-weighted MRI, the digastric anterior muscle demonstrated negative correlations in the submental left (coefficient: -0.45, 95% CI: -0.72 to -0.06). The genioglossus muscle exhibited a negative correlation in the intraoral left region (coefficient: -0.47, 95% CI: -0.74 to -0.07). The mylohyoid muscle showed negative correlations in the intraoral left (coefficient: -0.48, 95% CI: -0.75 to -0.09). Additionally, the tongue base demonstrated a negative correlation in the intraoral left region (coefficient: -0.42, 95% CI: -0.71 to -0.01). The Oral Tongue muscle did not show any notable correlations with p-values below 0.05. A negative correlation in the analysis indicating that as the muscle volume decreases, the HN-LEF score increases, as shown in Figure 2.

**Table III.** Association Between Muscle/Structure Volumes and Novel HN-LEF Scores in Submental and Intraoral Regions for T1- and T2-weighted MRI Sequences.

| Muscle/Structure | MRI Sequences |  | Submental Region Left | Submental Region Right | Intraoral Left | Intraoral Right |
| --- | --- | --- | --- | --- | --- | --- |
| Digastric Anterior | T1 | Coefficient (95% CI) | -0.41 (-0.65, -0.10) | -0.32 (-0.58, 0.01) | -0.09 (-0.40, 0.24) | -0.15 (-0.45, 0.18) |
|  |  | *p* | 0.01 † | 0.06 | 0.61 | 0.37 |
|  | T2 | Coefficient (95% CI) | -0.45 (-0.72, -0.06) | -0.40 (-0.69, -0.01) | -0.33 (-0.64, 0.07) | -0.33 (-0.64, 0.08) |
|  |  | *p* | 0.03 † | 0.05 ‡ | 0.11 | 0.11 |
| Genioglossus | T1 | Coefficient (95% CI) | -0.03 (-0.36, 0.33) | -0.03 (-0.36, 0.31) | -0.20 (-0.50, 0.14) | 0.16 (-0.18, 0.47) |
|  |  | *p* | 0.85 | 0.86 | 0.24 | 0.36 |
|  | T2 | Coefficient (95% CI) | 0.14 (-0.29, 0.52) | 0.12 (-0.30, 0.51) | -0.47 (-0.74, -0.07) | -0.12 (-0.51, 0.31) |
|  |  | *p* | 0.52 | 0.57 | 0.02 † | 0.59 |
| Geniohyoid | T1 | Coefficient (95% CI) | 0.11 (-0.25, 0.44) | 0.19 (-0.17, 0.51) | -0.15 (-0.47, 0.21) | 0.04 (-0.31, 0.38) |
|  |  | *p* | 0.56 | 0.29 | 0.41 | 0.82 |
|  | T2 | Coefficient (95% CI) | 0.36 (-0.10, 0.69) | 0.42 (-0.03, 0.72) | -0.20 (-0.60, 0.26) | 0.11 (-0.35, 0.53) |
|  |  | *p* | 0.12 | 0.07 | 0.38 | 0.65 |
| Mylohyoid | T1 | Coefficient (95% CI) | -0.36 (-0.62, 0.03) | -0.27 (-0.55, 0.07) | -0.30 (-0.58, 0.03) | -0.20 (-0.50, 0.15) |
|  |  | *p* | 0.04 † | 0.11 | 0.08 | 0.26 |
|  | T2 | Coefficient (95% CI) | -0.24(-0.57, 0.22) | -0.16(-0.52, 0.3) | -0.48(-0.75, -0.09) | -0.31(-0.64, -0.11) |
|  |  | *p* | 0.27 | 0.46 | 0.02 † | 0.14 |
| Tongue Base | T1 | Coefficient (95% CI) | 0.05 (-0.30, 0.38) | -0.00 (-0.35, 0.34) | -0.07 (-0.40, 0.28) | -0.09 (-0.42, 0.26) |
|  |  | *p* | 0.79 | 0.98 | 0.69 | 0.6 |
|  | T2 | Coefficient (95% CI) | 0.11 (-0.32, 0.5) | 0.12 (-0.3, 0.51) | -0.42 (-0.71, -0.01) | -0.04 (-0.45, 0.38) |
|  |  | *p* | 0.62 | 0.57 | 0.04 † | 0.85 |
| Oral Tongue | T1 | Coefficient (95% CI) | -0.02 (-0.37, 0.33) | -0.09 (-0.42, 0.27) | -0.37 (-0.63, 0.02) | 0.02 (-0.33, 0.37) |
|  |  | *p* | 0.91 | 0.63 | 0.05 ‡ | 0.9 |
|  | T2 | Coefficient (95% CI) | 0.23 (-0.20, 0.59) | 0.20 (-0.22, 0.57) | -0.19 (-0.56, 0.24) | 0.21 (-0.23, 0.57) |
|  |  | *p* | 0.29 | 0.35 | 0.39 | 0.35 |
| †: a p-value smaller than 0.05; ‡: a p-value equal to 0.05 |  |  |  |  |  |  |

**Figure 2.**
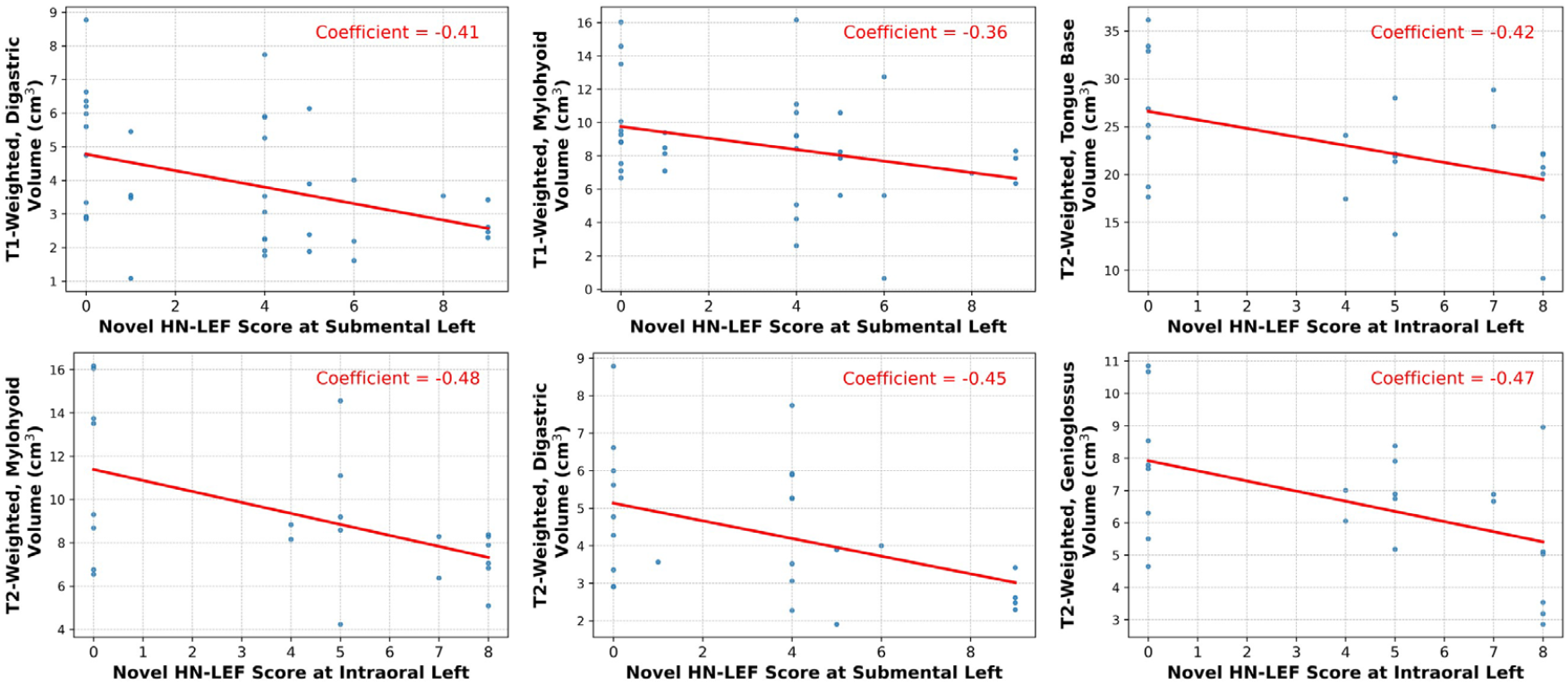
Linear correlations between the novel HN-LEF score and the structural volumes in the MRI imaging.

## 4. Discussion

In this study, we examined the correlation between palpation-based LEF assessments and MRI signal intensities across different HN-LEF categories in T1- and T2-weighted MRI images. While MRI stands as a powerful tool, some practical constraints can limit its utility as a frequent, first-line diagnostic tool. Conducting MRI requires specialized equipment and skilled personnel, making it costly and less accessible in some clinical settings. Additionally, MRI scans demand patients to remain still for long periods, which can be challenging for those with severe symptoms or discomfort. Furthermore, for HNC patients, the presence of metallic stabilizers used to support the mandible due to osteoradionecrosis can pose safety risks during MRI, as these materials are not MRI-safe (15).

The differential signal intensities observed in T1- and T2-weighted MRI images provide new insights into the structural and compositional differences among the HN-LEF categories. In T1 imaging, the heightened SI in the A-B category may reflect a higher density of cellular and extracellular components typical of mild fibrosis +/- edema. This could suggest that T1-weighted imaging is particularly sensitive to changes in tissue composition brought about by the initial stages of fibrosis mixed with edema. In T2-weighted imaging, the variation in signal intensity, particularly the highest intensity seen in category C followed by A-B compared to D and No-LEF, likely indicates different water content levels. The elevated signals in A-B and C suggest that these categories may retain more fluid, which is consistent with the presence of edema. The fact that category D shows lower intensity than No-LEF in T2 images is particularly noteworthy. It suggests that in cases of fibrosis only (category D), the tissue may become more dehydrated or denser, which reduces its ability to retain water, thus lowering the T2 signal intensity (which is counter to our hypothesis about T2). The distinct patterns observed in MRI signal intensities among the different LEF categories underscore the metric’s potential sensitivity to physiological changes within the tissue. This sensitivity highlights the utility of the LEF classification in providing diagnostic insights that are not only based on physical examination but are also corroborated by advanced imaging techniques. Thus, the data point toward validity of both metrics.

The observed negative correlations between the novel HN-LEF scores and the muscle volumes in T2-weighted MRI across various FOM muscle sites indicate a decrease in muscle volume with increasing severity of lymphedema and fibrosis. These correlations suggest a potential structural deterioration or atrophy in response to progressive LEF, prominently revealed through T2-weighted imaging. Notably, the relative scarcity of such correlations in T1-weighted images may highlight T2-weighted MRI’s superior sensitivity to the changes in water content and tissue composition associated with these conditions. This pattern suggests that as the severity and complexity of LEF, as encapsulated by the novel HN-LEF score, increase, there tends to be an overall decrease in the volume of affected muscles.

The findings of this study suggest that our novel proposed HN-LEF score holds potential as a biomarker for assessing lymphedema and fibrosis in HNC survivors. This potential arises from the ability of the LEF score to differentiate between various degrees and types of tissue changes associated with these conditions. By establishing a clear correlation between palpation-based assessments and MRI findings, the HN-LEF score could facilitate more accurate diagnoses and enable clinicians to monitor the progression or regression of LEF more effectively. This study, therefore, not only provides hypothesis-generating data about MRI biomarkers of HN-LEF, but also strengthens the evidence-base supporting palpation-based assessments in characterizing the late effect of the HNC treatment. Thus, the data reinforce the critical role of palpation-based HN-LEF grading in routine clinical evaluations.

### Limitations

This study encounters several limitations that should be considered. First, while the HN-LEF grading system has been applied with consensus between two clinicians, the long-term consistency and reliability of these assessments across different raters still need to be fully analyzed. Additionally, a challenge involves the alignment of the MRI and HN-LEF regions of interest (ROIs). Although we examined the signal intensity distribution within these ROIs, an exact match between the MRI ROIs and HN-LEF ROIs was not achieved in our statistical analysis, which may impact the accuracy of correlating MRI findings with clinical assessments. Moreover, the distribution of SI across different HN-LEF categories was not balanced, with varying numbers of measurements in each category, potentially affecting the statistical power and outcomes. Furthermore, the study is based on a relatively small dataset composed exclusively of patients exhibiting at least moderate fibrosis in some cervical regions. This limited sample size and the homogeneity in fibrosis severity could restrict the generalizability of our findings.

### Conclusion

This study provides preliminary investigation supporting the potential of MRI signal intensity as an imaging biomarker for assessing edematous and fibrotic soft tissue states in HNC patients. The observed inverse correlation between HN-LEF scores and muscle volumes suggests that as patients transition from edema to fibrosis, structural volume decreases, and points to potential value of a novel HN-LEF score from the validated HN-LEF classification system. Our findings underscore the potential for using imaging metrics to enhance the quantification of LEF, which could improve early detection and precise measurement of late effects in HNC survivors. Further research should focus on validating these findings and exploring their clinical applications.

## Data Availability

Data availability statement: In accordance with the Final NIH Policy for Data Management and Sharing, NOT-OD-21-013, anonymized tabular analytic and NIFTI data that support the findings of this study are openly available in an NIH-supported generalist scientific data repository (figshare) no later than the time of an associated peer-reviewed publication; while public data is embargoed pending peer review, the data is available upon request pre-peer-review through email to the corresponding author.

## References

1. Jackson LK, Ridner SH, Deng J, Bartow C, Mannion K, Niermann K, Gilbert J, Dietrich MS, Cmelak AJ, Murphy BA. Internal Lymphedema Correlates with Subjective and Objective Measures of Dysphagia in Head and Neck Cancer Patients. J Palliat Med. 2016;19(9):949–56. doi: 10.1089/jpm.2016.0018. PubMed PMID: 27227341; PMCID: PMC5011629.

2. Cohen EE, LaMonte SJ, Erb NL, Beckman KL, Sadeghi N, Hutcheson KA, Stubblefield MD, Abbott DM, Fisher PS, Stein KD, Lyman GH, Pratt-Chapman ML. American Cancer Society Head and Neck Cancer Survivorship Care Guideline. CA Cancer J Clin. 2016;66(3):203–39. doi: 10.3322/caac.21343. PubMed PMID: 27002678.

3. Ramia P, Bodgi L, Mahmoud D, Mohammad MA, Youssef B, Kopek N, Al-Shamsi H, Dagher M, Abu-Gheida I. Radiation-Induced Fibrosis in Patients with Head and Neck Cancer: A Review of Pathogenesis and Clinical Outcomes. Clin Med Insights-On. 2022;16. doi: Artn 11795549211036898 10.1177/11795549211036898. PubMed PMID: WOS:000751865000001.

4. Starmer H, Cherry MG, Patterson J, Young B, Fleming J. Assessment of Measures of Head and Neck Lymphedema Following Head and Neck Cancer Treatment: A Systematic Review. Lymphatic Research and Biology. 2023;21(1):42–51. doi: 10.1089/lrb.2021.0100. PubMed PMID: WOS:000808341700001.

5. Deng J, Murphy BA, Dietrich MS, Wells N, Wallston KA, Sinard RJ, Cmelak AJ, Gilbert J, Ridner SH. Impact of secondary lymphedema after head and neck cancer treatment on symptoms, functional status, and quality of life. Head Neck. 2013;35(7):1026–35. doi: 10.1002/hed.23084. PubMed PMID: 22791550; PMCID: 4017911.

6. Deng J, Dietrich MS, Aulino JM, Sinard RJ, Mannion K, Murphy BA. Longitudinal Pattern of Lymphedema and Fibrosis in Patients With Oral Cavity or Oropharyngeal Cancer: A Prospective Study. International Journal of Radiation Oncology* Biology* Physics. 2023.

7. Burgos-Mansilla B, Galiano-Castillo N, Lozano-Lozano M, Fernández-Lao C, Lopez-Garzon M, Arroyo-Morales M. Effect of physical therapy modalities on quality of life of head and neck cancer survivors: a systematic review with meta-analysis. J Clin Med. 2021;10(20):4696.

8. Deng J, Dietrich MS, Ridner SH, Fleischer AC, Wells N, Murphy BA. Preliminary evaluation of reliability and validity of head and neck external lymphedema and fibrosis assessment criteria. Eur J Oncol Nurs. 2016;22:63–70. Epub 2016/05/18. doi: 10.1016/j.ejon.2016.02.001. PubMed PMID: 27179894.

9. Purcell A, Nixon J, Fleming J, McCann A, Porceddu S. Measuring head and neck lymphedema: The “ALOHA’’ trial. Head Neck-J Sci Spec. 2016;38(1):79–84. doi: 10.1002/hed.23853. PubMed PMID: WOS:000368737600019.

10. Fadhil M, Singh R, Havas T, Jacobson I. Systematic review of head and neck lymphedema assessment. Head Neck-J Sci Spec. 2022;44(10):2301–15. doi: 10.1002/hed.27136. PubMed PMID: WOS:000822976700001.

11. Deng J, Ridner SH, Dietrich MS, Wells N, Murphy BA. Assessment of External Lymphedema in Patients With Head and Neck Cancer: A Comparison of Four Scales. Oncology nursing forum. 2013;40(5):501–6. doi: 10.1188/13.Onf.501-506. PubMed PMID: WOS:000324565300013.

12. Deng J, Ridner SH, Wells N, Dietrich MS, Murphy BA. Development and preliminary testing of head and neck cancer related external lymphedema and fibrosis assessment criteria. Eur J Oncol Nurs. 2015;19(1):75–80. Epub 2014/09/06. doi: 10.1016/j.ejon.2014.07.006. PubMed PMID: 25190635.

13. Taylor AJ, Salerno M, Dharmakumar R, Jerosch-Herold M. T1 Mapping: Basic Techniques and Clinical Applications. JACC Cardiovascular imaging. 2016;9(1):67–81. Epub 2016/01/15. doi: 10.1016/j.jcmg.2015.11.005. PubMed PMID: 26762877.

14. Arrivé L, Derhy S, Dlimi C, El Mouhadi S, Monnier-Cholley L, Becker C. Noncontrast Magnetic Resonance Lymphography for Evaluation of Lymph Node Transfer for Secondary Upper Limb Lymphedema. Plastic and reconstructive surgery. 2017;140(6):806e–11e. doi: 10.1097/Prs.0000000000003862. PubMed PMID: WOS:000418597700005.

15. Cellina M, Gibelli D, Martinenghi C, Giardini D, Soresina M, Menozzi A, Oliva G, Carrafiello G. Non-contrast magnetic resonance lymphography (NCMRL) in cancer-related secondary lymphedema: acquisition technique and imaging findings. Radiologia Medica. 2021;126(11):1477–86. doi: 10.1007/s11547-021-01410-3. PubMed PMID: WOS:000684080600001.

16. Salehi BP, Sibley RC, Friedman R, Kim G, Singhal D, Loening AM, Tsai LL. MRI of Lymphedema. J Magn Reson Imaging. 2023;57(4):977–91. doi: 10.1002/jmri.28496. PubMed PMID: WOS:000871006400001.

17. Hutcheson K, McMillan H, Warneke C, Porsche C, Savage K, Buoy S, Wang JH, Woodman K, Lai S, Fuller C. Manual Therapy for Fibrosis-Related Late Effect Dysphagia in head and neck cancer survivors: the pilot MANTLE trial. BMJ open. 2021;11(8). doi: ARTN e047830 10.1136/bmjopen-2020-047830. PubMed PMID: WOS:000692198100004.

18. McMillan H, Barbon CEA, Cardoso R, Sedory A, Buoy S, Porsche C, Savage K, Mayo L, Hutcheson KA. Manual Therapy for Patients With Radiation-Associated Trismus After Head and Neck Cancer. Jama Otolaryngol. 2022;148(5):418–25. doi: 10.1001/jamaoto.2022.0082. PubMed PMID: WOS:000770396800002.

19. Hutcheson KA, Barrow MP, Barringer DA, Knott JK, Lin HY, Weber RS, Fuller CD, Lai SY, Alvarez CP, Raut J, Lazarus CL, May A, Patterson J, Roe JWG, Starmer HM, Lewin JS. Dynamic Imaging Grade of Swallowing Toxicity (DIGEST): Scale Development and Validation. Cancer-Am Cancer Soc. 2017;123(1):62–70. doi: 10.1002/cncr.30283. PubMed PMID: WOS:000394719100009.

20. Hutcheson KA, Barbon CEA, Alvarez CP, Warneke CL. Refining measurement of swallowing safety in the Dynamic Imaging Grade of Swallowing Toxicity (DIGEST) criteria: Validation of DIGEST version 2. Cancer-Am Cancer Soc. 2022;128(7):1458–66. doi: 10.1002/cncr.34079. PubMed PMID: WOS:000738970600001.

21. Chen AP, Setser A, Anadkat MJ, Cotliar J, Olsen EA, Garden BC, Lacouture ME. Grading dermatologic adverse events of cancer treatments: the Common Terminology Criteria for Adverse Events Version 4.0. Journal of the American Academy of Dermatology. 2012;67(5):1025–39.

